# A clinical audit of postoperative care after emergency caesarean section for obstructed labour at a national referral hospital in Uganda

**DOI:** 10.64898/2026.10.21.26366170

**Authors:** Mwanje Derrick, Ononge Sam, Nanzira Racheal Samantha, Byaruhanga Kayogoza Emmanuel, Igirimbabazi Ssebuddo Pious, Kagawa Nantamu Mike

## Abstract

**Background:** Postoperative complications after emergency caesarean section (C/S) due to obstructed labour (O/L) are a significant contributing factor to maternal morbidity in low-resource settings. At Kawempe National Referral Hospital (KNRH) in Kampala, Uganda, hospital records from late 2024 showed an overall adverse outcome rate of approximately 10% among all women delivered by caesarean section, with postpartum haemorrhage (PPH) and surgical site infection (SSI) among the recorded outcomes. Women who deliver by C/S for O/L are particularly a high-risk group, yet adherence to postoperative care standards in this group had not been systematically evaluated. This audit therefore aimed at assessing whether a clinical audit could improve the quality of postoperative care among women undergoing C/S for O/L.

**Methods:** A criterion-based audit cycle was conducted at a national referral hospital in Kampala, Uganda over six months. At baseline, 150 case files of mothers who delivered by caesarean section due to O/L were reviewed retrospectively, and care practices were evaluated against six care standards derived from World Health Organization (WHO) and Médecins Sans Frontieres (MSF) guidelines. These included: monitoring of vital signs (blood pressure, pulse rate, respiratory rate, temperature, oxygen saturation), administration of analgesia, surgical site care, administration of prophylactic and empirical antibiotics, monitoring for postpartum haemorrhage (monitoring uterine tone and vaginal bleeding), and bladder care. Findings were shared with stakeholders, and gaps were identified. Interventions such as continuous medical education (CME) sessions, protocol dissemination, and workflow adjustments, co-developed with stakeholders, were rigorously implemented for 2 months and 2 weeks. An endline audit was done by reviewing 149 case files. Data analysis involved a comparative description of the baseline and endline audits to determine statistical significance using a chi-square test. To assess the overall level of adherence to postoperative care standards, a composite indicator overall mean care score was computed for both baseline and endline phases. This metric was derived by aggregating compliance across all evaluated components of the standards.

**Results:** Statistically significant improvements were noted in three standards: Antibiotic administration-prophylactic antibiotics administration (38.0% to 60.4%; p<0.001) and empiric antibiotics administration on Day 1(metronidazole-13.7% to 96.2%, p<0.001), urine output monitoring on Day 1(17.3% to 28.9%; p=0.012), and analgesia administration (paracetamol and rectal diclofenac-50% to above 80%, p<0.002). However, critical gaps persisted, with no observed improvement in vital sign monitoring and surgical site care, and a paradoxical decline in uterine tone monitoring (90.0 to 88.7%; p=0.667). The overall mean care score improved from 46.7% at baseline to 61.1% at endline (absolute increase of +14.4% points).

**Conclusion:** The results of this study show that a clinical audit is a feasible quality improvement tool that can enhance postoperative care for mothers who deliver by C/S due to O/L in low-resource settings. However, there is a need to evaluate the other standards that did not have an improvement.

## Introduction

Maternal mortality and morbidity remain major public health challenges worldwide, with the greatest burden occurring in low- and middle-income countries. Despite substantial progress in maternal healthcare over the past two decades, an estimated 287,000 women died from pregnancy- and childbirth-related causes in 2020, with approximately 70% of these deaths occurring in sub-Saharan Africa [1]. Obstructed labour remains one of the leading direct causes of maternal and perinatal morbidity and mortality in the region, contributing to complications such as postpartum haemorrhage, puerperal sepsis, uterine rupture, obstetric fistula, and perinatal death [2]. Timely access to comprehensive emergency obstetric care, including caesarean section, is therefore essential for preventing adverse maternal and neonatal outcomes.

Caesarean section is among the most frequently performed surgical procedures worldwide and plays a critical role in managing obstetric emergencies. The global caesarean section rate increased from approximately 7% in 1990 to more than 21% in 2021, reflecting increased access to surgical obstetric care in many settings[3]. While caesarean section can be lifesaving for both mother and baby, it is associated with a higher risk of postoperative complications compared with vaginal delivery. Women undergoing emergency caesarean section, particularly following obstructed labour, are at increased risk of postpartum haemorrhage, surgical site infection, severe postoperative pain, thromboembolic events, prolonged hospitalization, and maternal death [4]. The risk of these complications is often greater in low-resource settings where healthcare systems face challenges related to staffing, infrastructure, supplies, and adherence to clinical guidelines.

Evidence suggests that a substantial proportion of postoperative morbidity following caesarean section can be prevented through adherence to recommended postoperative management practices. International and national guidelines recommend routine monitoring of vital signs, adequate pain control, timely administration of prophylactic antibiotics, wound assessment, bladder care, early mobilization, breastfeeding support, patient education, and discharge planning as key components of quality postoperative care [5]. Studies conducted in sub-Saharan Africa have reported that inadequate adherence to these standards is associated with increased rates of postpartum haemorrhage, surgical site infections, prolonged hospital stay, and other adverse maternal outcomes [6], [7]. Strengthening postoperative care therefore represents an important opportunity for reducing maternal morbidity following emergency caesarean section.

Clinical audit is a recognized quality improvement strategy that involves measuring current practice against predefined standards, identifying gaps in care, implementing corrective interventions, and re-evaluating performance. Previous obstetric audits conducted in low-resource settings have demonstrated improvements in adherence to evidence-based practices and maternal health outcomes following implementation of audit recommendations [8]. In Uganda, clinical audits have contributed to improvements in the management of maternal and neonatal conditions, including obstructed labour and preterm birth [8], [9]. However, there is limited evidence regarding adherence to postoperative care standards among women undergoing emergency caesarean section due to obstructed labour, particularly in high-volume maternity facilities.

Kawempe National Referral Hospital is one of Uganda’s largest maternity hospitals and manages a substantial number of women requiring emergency caesarean section for obstructed labour. Although these women are at increased risk of postoperative complications, the quality of postoperative care provided and the effect of clinical audit on adherence to recommended standards have not been adequately evaluated. This study therefore aimed to assess adherence to established postoperative management standards among mothers undergoing emergency caesarean section due to obstructed labour at Kawempe National Referral Hospital, implement targeted quality improvement interventions, and evaluate whether these interventions improved the quality of postoperative care.

## Materials and methods

### Study design

This was a criterion-based clinical audit conducted using a before-and-after design to assess adherence to recommended postoperative management standards among mothers who underwent emergency caesarean section due to obstructed labour at Kawempe National Referral Hospital (KNRH) in Kampala, Uganda. The hospital serves as a national referral center for obstetric, gynecological, neonatal, and reproductive health services and also functions as a teaching hospital for undergraduate and postgraduate health professional training. The hospital conducts a high volume of deliveries annually and receives referrals from health facilities within Kampala Metropolitan Area and other parts of Uganda.

The audit involved three main phases: a baseline audit to assess current practice against predefined standards, implementation of quality improvement interventions to address identified gaps, and an endline audit to determine whether adherence to the standards improved following implementation of the interventions. The audit cycle is presented in Fig **1**

**Fig 1.**
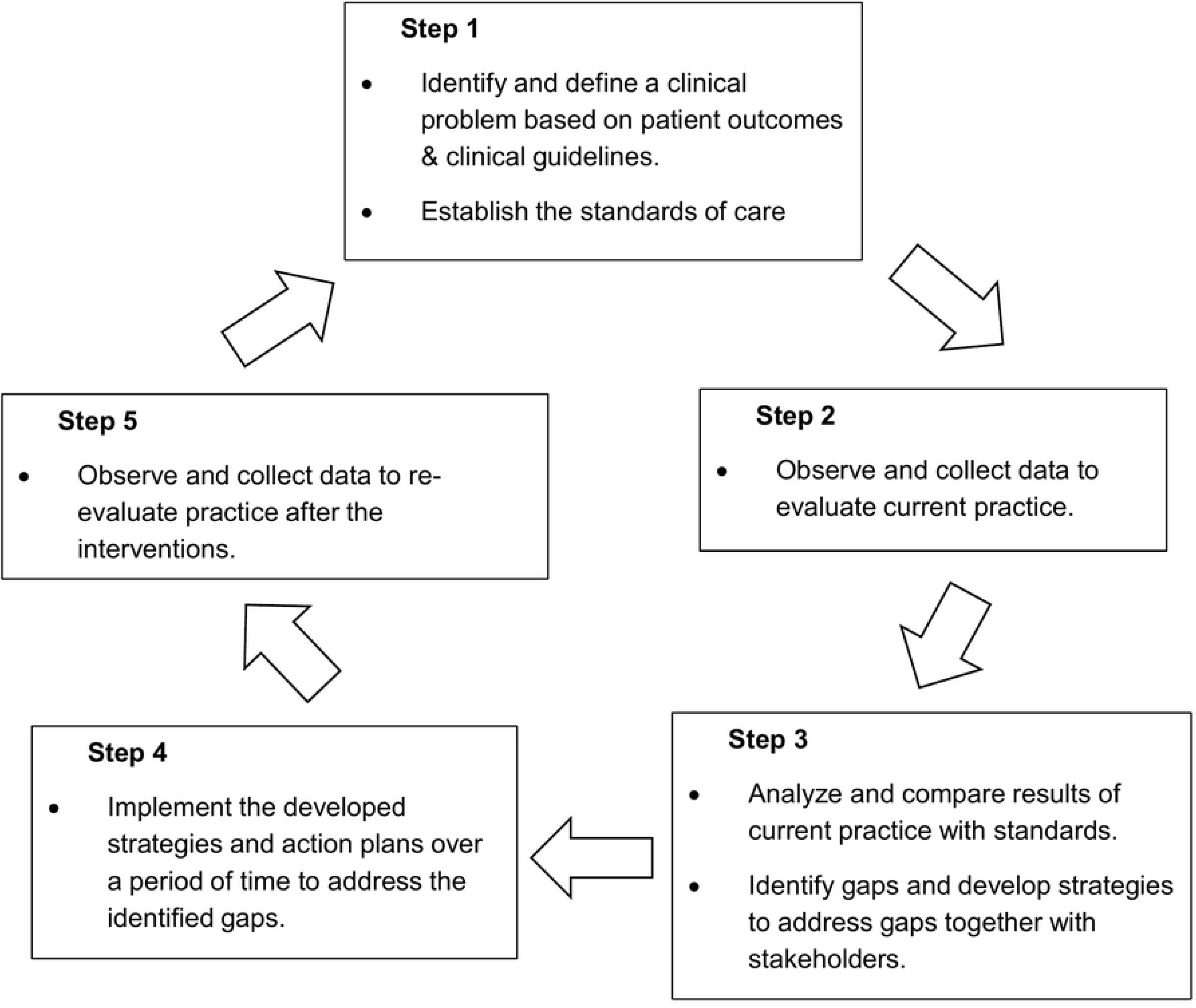
The audit cycle. Author-designed figure based on the principles of best practice in clinical audit outlined by the National Institute for Health and Care excellence (NICE) in 2002.

**Step 1:** Reviewed guidelines for the management of mothers after caesarean section due to obstructed labour to establish the standards. The standards used in this study, outlined in Table 1, were established by reviewing current guidelines by WHO-Integrated Management of Pregnancy and Childbirth: Managing Complications in Pregnancy and Childbirth, a guide for midwives and doctors [10]. The principles outlined in section 1, under “Postoperative Care Principles” (pages C-71 to C-75) were adopted. The standard on bladder care was derived from Médecins Sans Frontieres (MSF): Essential Obstetrics and Newborn Care-Practical guide for midwives and doctors with obstetric training and health care professionals who deal with obstetric emergencies [11].

**Table 1.** The six standards, identified gaps, reasons for non-compliance and recommended interventions.

| Standard | Gaps after baseline audit | Reason | Intervention/recommendation |
| --- | --- | --- | --- |
| **Vital sign monitoring:**<br>1/4hrly for 1hr, then 1/2hrly for 2hrs, then 1hrly for 6hrs, then twice or thrice a day on Day 1 and Day 2 | -Inadequate vital sign monitoring especially in the first critical postoperative hours, i.e. 1hr, 2hrs and 6hr intervals. | -Lack of knowledge on the standards.<br>-Inadequate documentation of measured parameters.<br>-Faulty monitoring machines. | -CME sessions (3) on standard postoperative care.<br>-Timely documentation of all given care in the hospital information system.<br>-In-charge on postnatal ward to keep reminding the team through their social media platform (WhatsApp) and to do spot checks on how documentation is being done once a week.<br>-In-charge to ask the biomedical team to repair the faulty monitoring machines. |
| **Uterine tone & vaginal bleeding monitoring:**<br>Every 15-30mins in first 3hrs, every 1hr in the next 6hrs then once or twice a day on Day 1 and Day 2. | -Standard frequency of examination not observed especially in the first critical hours after surgery i.e. 1hr, 2hrs and 6hrs intervals. | - Large patient to caregiver ratio.<br>-Lack of knowledge on standards.<br>-Poor documentation.<br>-Lack of adequate training on hospital information system use. | -Administration to recruit more healthcare providers.<br>-In-charge to adjust the rotas so that busy days have more people on duty.<br>-CME sessions on standard postoperative care.<br>-IT team to train clinical staff on all aspects of the hospital information system and to address the slow internet challenge. |
| **Surgical site care:**<br>Uncover wound on day 1 to inspect the site. Only change dressing if dirty or soaked. | -Wounds were never examined in 5.3% of mothers | - Intern doctors pay no attention to wound care during ward rounds.<br>-Lack of knowledge on standard surgical site care. | - CME sessions (3) on standard surgical site care.<br>-In-charge to brief intern doctors before they start ward rounds so they examine patient surgical sites and to document in the hospital information system. |
| **Bladder care.**<br>For mothers with high risk for bladder injury like in severe obstruction, leave catheter for 14 days. | -Premature removal of urine catheters in mothers where it was prescribed for 14 days | -Duration of catheter is never clearly indicated on the file.<br>-Failure to refer to the surgeon's postoperative instructions concerning catheter duration. | -Surgeons to indicate on top of treatment files and in the postoperative instructions the duration of the urethral catheter.<br>-Surgeons to counsel mothers/caretakers on duration of urethral catheter and when it should be removed. |
| **Prophylactic & empiric antibiotic therapy.**<br>Prophylaxis should be given 15- 60 minutes before incision.<br>Repeat parenteral doses for 48 hrs. | -Up to 62% of mothers did not get prophylactic antibiotics despite availability of drugs<br>- Poor adherence to prescribed frequency and duration of empiric antibiotics. | -High patient to caregiver ratio so timely administration of antibiotics is missed in some patients.<br>-Inadequate documentation when drugs are administered. | -In-charge of anaesthesia to ensure that busy days have a minimum of two anaesthetists on duty.<br>-Administration of prophylactic antibiotics immediately the mother enters theatre before anesthesia administration.<br>-All empiric antibiotic doses of the day to be picked at once from the pharmacy by the caretakers.<br>-Nurses on postnatal ward to document administered drugs immediately using the mobile laptops. |
| **Analgesia:**<br>Should be multimodal.<br>Parenteral route in the first 24hours.<br>Oral route analgesia after day 1. | -Poor adherence to prescribed frequency and duration. | -Inadequate documentation when drugs are administered.<br>-Occasional stock outs of prescribed analgesics. | - Caretakers to pick all analgesia doses of the day at once from the pharmacy.<br>-Nurses to document administered drugs immediately using the mobile laptops.<br>-Self-administered analgesia options like rectal diclofenac should be prioritized.<br>-Encourage mothers/caretakers to remind nurses about subsequent doses. |

**Step 2:** Baseline audit. A total of 150 case files of eligible mothers on the postnatal ward were reviewed against a standardized checklist at the time of discharge over a period 6 weeks from November 24^th^, 2025 to January 6^th^, 2026. Two trained midwife research assistants, not stationed at the postnatal ward, collected data to minimise bias. A specialist obstetrician provided expert guidance. Where catheter-related documentation was missing, mothers were interviewed at discharge following informed consent, to establish presence or absence of the urethral catheter if it had been prescribed for 14 days.

**Step 3:** Analysis and feedback: This phase lasted 3 weeks. The data collected from the baseline audit was analyzed to identify patterns, trends, and areas of non-compliance with the established standard guidelines. It was a comparison between the standard practices and the actual care provided, highlighting areas where care did not meet the expected criteria. The findings were shared with key stakeholders, including obstetricians, anesthesiologists, nursing staff, and hospital management. Root causes of gaps were identified, and recommendations and improvement strategies were collaboratively developed.

**Step 4:** Several recommendations (Table 1) were rigorously implemented over a period of 2 months and 2 weeks from January 30^th^, 2026 to April 14^th^, 2026, by both the study team and the clinical team: Three CME sessions were conducted about standard postoperative care. Postoperative care protocols were displayed in clinical areas where they were easily visible and and workflow adjustments were implemented.

**Step 5:** Endline audit. A re-evaluation of practice after implementing the recommendations involved a review of 149 files of eligible mothers over a period of 5 weeks, from April 16^th^, 2026 to May 20^th^, 2026, against the same checklist used in step 2. The aim was to determine whether the care provided after implementing the recommendations aligned with the standards.

### Sample size and sampling procedure

Used the Kish Leslie formula of 1965 for sample size estimation for two tailed-test i.e. a baseline visit, intervention and then an endline visit. 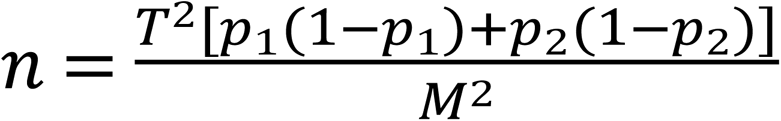

Where **n** is the required sample size, **T** is the confidence level at 95% (standard value 1.96), **M** is the margin of error at 5% (standard value of 0.05), **p_1_** and **p_2_** are the proportions of the mean care score for the baseline and endline audits respectively. **p_1_** being 80% and **p_2_** being 98.2% for one of the standards (vital sign monitoring) in a criterion based audit done in Malawi [12].

Computing in the formula above gave a total sample size **n** of 273.024 (274) for the two audits.

In order to improve the power of the study, a total sample size of 299 was used, 150 for the baseline audit and 149 for the endline audit.

All eligible mothers who underwent emergency caesarean section due to obstructed labour during the audit periods were consecutively enrolled until the above sample size was attained. Consecutive sampling was chosen to minimize selection bias and ensure inclusion of all eligible participants presenting during the study period.

### Data collection procedures

Data were collected using a structured audit checklist developed from the predefined audit standards (Table 1). Information was obtained from patient medical records, nursing charts, medication administration records, postoperative monitoring charts, and direct observation where applicable.

During the baseline audit, data were collected on adherence to each postoperative management standard as illustrated in the audit cycle ( Fig **1**). The findings were presented to relevant stakeholders, including obstetricians, medical officers, midwives, anesthetists, and hospital administrators. The same process was repeated in the endline audit after implementing the recommendations.

### Statistical methods

Data were entered, cleaned, and analyzed using STATA version 16.0 (StataCorp, College Station, TX, USA). Descriptive statistics were used to summarize study findings. Categorical variables were summarized using frequencies and percentages. A comparative analysis was conducted between the baseline audit and endline audit to assess changes in adherence to each postoperative management standard. For each audit criterion, the proportion of mothers receiving the recommended standards of care were compared between baseline and endline audit. The primary measure of effect was the risk difference (RD), calculated as the difference in compliance proportions between the endline audit and baseline audit. Risk differences were presented together with corresponding 95% confidence intervals (95% CI). Statistical significance was assessed using two-sample tests of proportions, and p-values were reported. A p-value less than 0.05 was considered statistically significant.

To assess the overall level of adherence to postoperative care standards, a composite indicator overall mean care score was computed for both baseline and endline phases. This metric was derived by aggregating compliance across all evaluated components of the standards.

Each component of the standards was operationalized as a binary or proportion-based compliance indicator and expressed as a percentage compliance. All components were treated with equal weight.

The overall mean care score was then calculated by summing the percentage compliance values for all assessed components and dividing by the total number of components of standards evaluated.

Overall Mean Care Score = (Sum of percentage compliance across all components of standards) / (Total number of components of standards)

A total of 65 components were included in the analysis, spanning vital sign monitoring, surgical site care, abdominal assessment, vaginal bleeding monitoring, bladder care, antibiotic and analgesics administration practices.

### Ethical considerations

Clearance was granted by the Department of Obstetrics and Gynecology. Ethical approval, including a waiver of consent for medical records review, was granted by the School of Medicine Research Ethics Committee (Mak-SOMREC-2025-599). Administrative clearance was granted by Kawempe National Referral Hospital and informed consent was obtained from respondents after explaining the purpose of the study. Medical records access commenced on November 24^th^, 2025 and was concluded on May 20^th^, 2026. During data collection, the research team could access potentially identifiable information, including participant’s names and inpatient numbers; however, all data were anonymized prior to analysis.

## Results

### Baseline and endline audits

A total of 299 files were reviewed. 150 files in the baseline audit and 149 files in the endline audit. The study was conducted over a period of 6 months from November 2025 to May 2026. The baseline audit was conducted over six weeks, from November 24^th^, 2025 to January 6^th^, 2026. This was followed by three weeks of data analysis and dissemination of findings, then two months and two weeks of implementing recommended interventions, from January 30^th^ to April 14^th^, 2026, and five weeks for the endline audit, from April 16^th^ to May 20^th^, 2026.

#### Demographic and clinical characteristics of study participants

The findings in Table 2 demonstrate some shifts in the demographic and clinical profiles of mothers diagnosed with O/L between the baseline and the endline audits. The proportion of women diagnosed with O/L at admission increased slightly from 36.4% to 38.3%. All mothers (100%) in both phases had a diagnosis of O/L prior to surgery. The proportion of decisions made by specialists increased from 6.6% to 14.1%.

**Table 2.** Demographic and clinical profile of mothers (Baseline vs Endline)

| Variable | Baseline |  | Endline |  |
| --- | --- | --- | --- | --- |
| Gravidity | Frequency (n=150) | Percentage (%) | Frequency (n=149) | Percentage (%) |
| Primigravida | 75 | 50.3% | 62 | 41.6% |
| Multigravida | 60 | 39.7% | 75 | 50.3% |
| Grand multigravida | 15 | 9.9% | 12 | 8.1% |
| **Diagnosis at admission** |  |  |  |  |
| Active labour | 38 | 25.2% | 39 | 26.2% |
| Latent labour | 57 | 38.4% | 53 | 35.6% |
| Obstructed labour | 55 | 36.4% | 57 | 38.3% |
| **Final diagnosis prior to caesarean delivery** |  |  |  |  |
| Obstructed labour | 150 | 100.0% | 149 | 100.0% |
| **Cadre sanctioning caesarean delivery** |  |  |  |  |
| Resident | 140 | 93.4% | 128 | 85.9% |
| Specialist | 10 | 6.6% | 21 | 14.1% |
| **Cadre that performed the surgery** |  |  |  |  |
| Resident | 150 | 100.0% | 149 | 100.0% |

#### Vital sign monitoring

There was a marginal increase in blood pressure monitoring during the first postoperative hour (Risk difference (RD) = +2.0%), although this change was not statistically significant. At the two-hour and six-hour intervals, there was no improvement (RD = 0%), indicating persistent non-adherence to recommended monitoring guidelines. Pulse rate monitoring followed a similar trend, with only a negligible improvement in the first hour (RD = +0.7%) and no change at subsequent time points.

Despite respiratory rate and temperature being part of the vital signs to be monitored during postoperative care, none of the participants had them monitored as per the standards in both audits.

#### Abdominal palpation for uterine tone and vaginal bleeding monitoring

There was no significant improvement in abdominal palpation for uterine tone and monitoring of vaginal bleeding (RD = −1.3% respectively). Standard was met on Day 1 and Day 2 in 90.0% of the mothers in the baseline. It however declined to 88.7% in the endline, but not statistically significant.

#### Surgical site care

A statistically significant decline in surgical site inspection was observed (RD = −8.1%). There was a slight reduction in the proportion of dirty dressings (RD = −2.0%), although this was not statistically significant.

#### Bladder care

Urine monitoring (Table 3) showed no change on Day 0 (RD = −0.6%), but a statistically significant improvement was observed on Day 1 (RD = +11.6%). This suggests that the intervention had a delayed effect, with improvements occurring during ward-based care rather than immediately postoperatively. On Day 2, the improvement was smaller and not statistically significant (RD = +2.8%). There was a modest, non-significant increase in 14-day catheter prescription (RD = +5.4%) and catheter presence at discharge (RD = +2.2%), suggesting a persistence in poor adherence to guidelines.

**Table 3.** Bladder care.

| Variable | Baseline<br>(n=150) | Endline<br>(n=149) | RD | 95% CI | P-Value |
| --- | --- | --- | --- | --- | --- |
| Urine colour and volume inspected as per review notes (Day 0) | 12 (8.0%) | 11 (7.4%) | -0.6 | (-4.8, +3.6) | 0.783 |
| Urine colour and volume inspected as per review notes (Day 1) | 26 (17.3%) | 43 (28.9%) | +11.6 | (+2.6, +20.6) | **0.012\*** |
| Urine colour and volume inspected as per review notes (Day 2) | 25 (16.7%) | 29 (19.5%) | +2.8 | (-5.8, +11.4) | 0.520 |
| Urine catheter prescribed for 2 weeks. | 74 (49.3%) | 81 (54.4%) | +5.4 | (-5.9, +16.9) | 0.347 |
| Urine catheter in-situ at discharge. (*Baseline: n=74, Endline: n=81*) | 24 (32.4%) | 28 (34.6%) | +2.2 | (-12.0, +16.4) | 0.758 |

RD, risk difference (endline proportion minus baseline proportion), expressed in percentage points; 95% CI, 95% confidence interval; \* Significant at p-value < 0.05.

#### Compliance to prophylaxis and empiric antibiotics administration

A statistically significant improvement was observed in prophylactic antibiotic administration from 38.0% to 60.4% (RD=+22.4, p-value <0.001.) Parenteral empiric antibiotic compliance remained high in both audit phases. Metronidazole compliance improved remarkably across all time points and parameters. Ceftriaxone showed significant improvement on Day 0 but these gains were not sustained on Day 1 and 2. Table 4 presents empiric antibiotic administration data at baseline and endline.

**Table 4.**
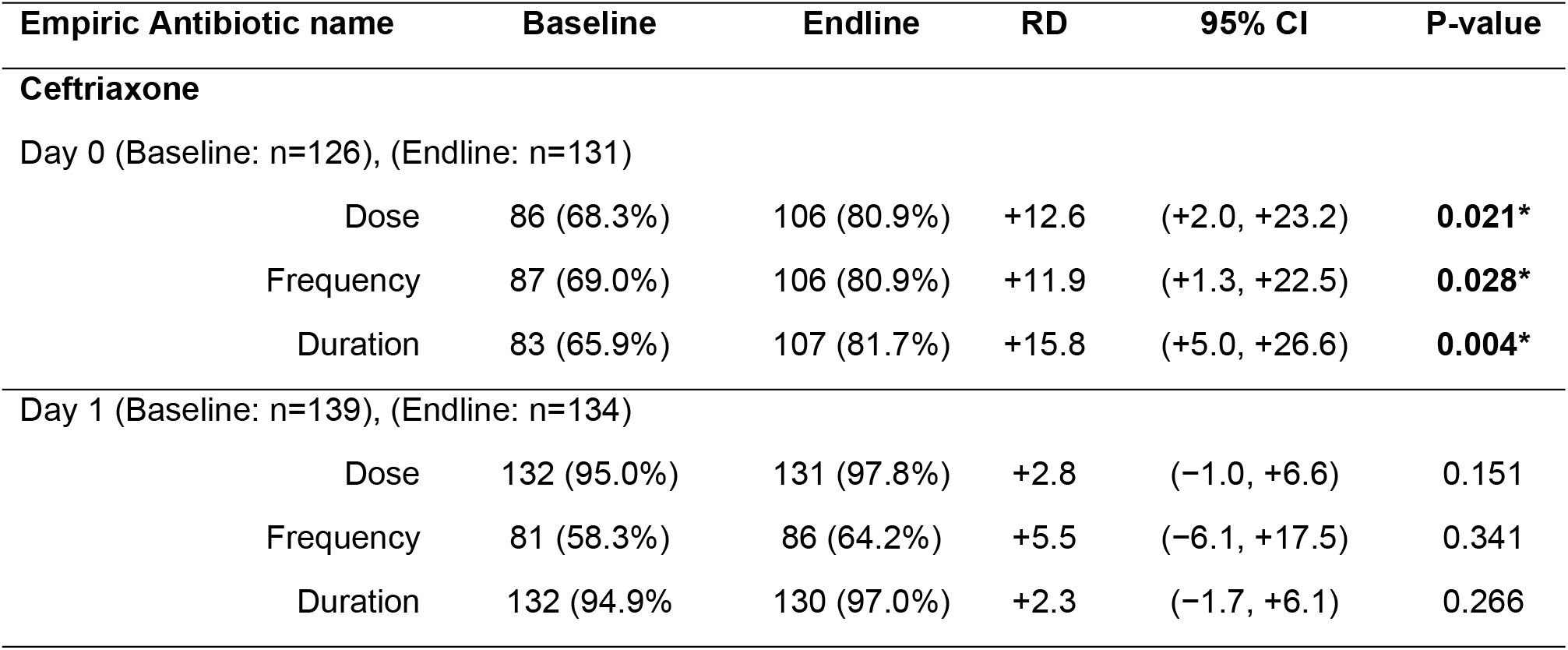
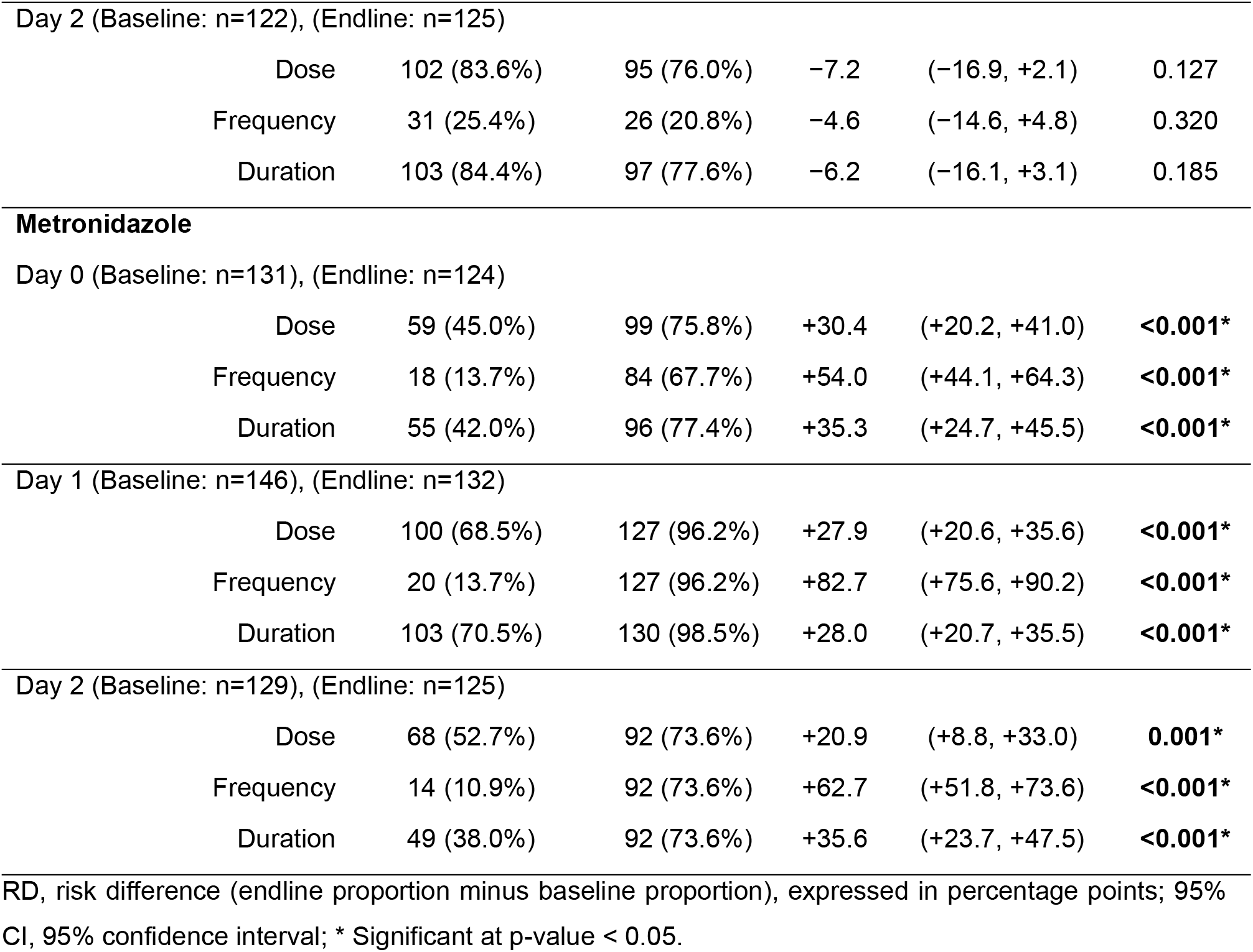
Compliance to empiric antibiotics prescription.

| Empiric Antibiotic name | Baseline | Endline | RD | 95% CI | P-value |
| --- | --- | --- | --- | --- | --- |
| **Ceftriaxone** |  |  |  |  |  |
| Day 0 (Baseline: n=126), (Endline: n=131) |  |  |  |  |  |
| Dose | 86 (68.3%) | 106 (80.9%) | +12.6 | (+2.0, +23.2) | **0.021\*** |
| Frequency | 87 (69.0%) | 106 (80.9%) | +11.9 | (+1.3, +22.5) | **0.028\*** |
| Duration | 83 (65.9%) | 107 (81.7%) | +15.8 | (+5.0, +26.6) | **0.004\*** |
| Day 1 (Baseline: n=139), (Endline: n=134) |  |  |  |  |  |
| Dose | 132 (95.0%) | 131 (97.8%) | +2.8 | (-1.0, +6.6) | 0.151 |
| Frequency | 81 (58.3%) | 86 (64.2%) | +5.5 | (-6.1, +17.5) | 0.341 |
| Duration | 132 (94.9%) | 130 (97.0%) | +2.3 | (-1.7, +6.1) | 0.266 |
| Day 2 (Baseline: n=122), (Endline: n=125) |  |  |  |  |  |
| Dose | 102 (83.6%) | 95 (76.0%) | -7.2 | (-16.9, +2.1) | 0.127 |
| Frequency | 31 (25.4%) | 26 (20.8%) | -4.6 | (-14.6, +4.8) | 0.320 |
| Duration | 103 (84.4%) | 97 (77.6%) | -6.2 | (-16.1, +3.1) | 0.185 |
| **Metronidazole** |  |  |  |  |  |
| Day 0 (Baseline: n=131), (Endline: n=124) |  |  |  |  |  |
| Dose | 59 (45.0%) | 99 (75.8%) | +30.4 | (+20.2, +41.0) | <0.001* |
| Frequency | 18 (13.7%) | 84 (67.7%) | +54.0 | (+44.1, +64.3) | <0.001* |
| Duration | 55 (42.0%) | 96 (77.4%) | +35.3 | (+24.7, +45.5) | <0.001* |
| Day 1 (Baseline: n=146), (Endline: n=132) |  |  |  |  |  |
| Dose | 100 (68.5%) | 127 (96.2%) | +27.9 | (+20.6, +35.6) | <0.001* |
| Frequency | 20 (13.7%) | 127 (96.2%) | +82.7 | (+75.6, +90.2) | <0.001* |
| Duration | 103 (70.5%) | 130 (98.5%) | +28.0 | (+20.7, +35.5) | <0.001* |
| Day 2 (Baseline: n=129), (Endline: n=125) |  |  |  |  |  |
| Dose | 68 (52.7%) | 92 (73.6%) | +20.9 | (+8.8, +33.0) | 0.001* |
| Frequency | 14 (10.9%) | 92 (73.6%) | +62.7 | (+51.8, +73.6) | <0.001* |
| Duration | 49 (38.0%) | 92 (73.6%) | +35.6 | (+23.7, +47.5) | <0.001* |

RD, risk difference (endline proportion minus baseline proportion), expressed in percentage points; 95% CI, 95% confidence interval; \* Significant at p-value < 0.05.

#### Compliance to analgesia prescription

Diclofenac showed statistically significant improvements in frequency and duration across all time points. Paracetamol showed strong improvements in dose and frequency, particularly on Day 1 and Day 2. Table 5 presents compliance to analgesia administration at baseline and endline.

**Table 5.** Analgesics compliance.

| Analgesic name | Baseline | Endline | RD | 95% CI | P-value |
| --- | --- | --- | --- | --- | --- |
| **Diclofenac** |  |  |  |  |  |
| Day 0 (Baseline: n=26), (Endline: n=28) |  |  |  |  |  |
| Dose | 13 (50.0%) | 24 (85.7%) | +35.7 | (+13.2, +60.1) | 0.002* |
| Frequency | 12 (46.2%) | 26 (92.8%) | +46.6 | (+21.4, +70.8) | <0.001* |
| Duration | 13 (50.0%) | 22 (78.6%) | +28.6 | (+9.7, +54.2) | **0.006\*** |
| Day 1 (Baseline: n=26), (Endline: n=25) |  |  |  |  |  |
| Dose | 12 (46.2%) | 20 (80.0%) | +33.8 | (+10.1, +59.1) | **0.006\*** |
| Frequency | 14 (53.8%) | 18 (72.0%) | +18.2 | (-7.3, +45.9) | 0.154 |
| Duration | 13 (50.0%) | 21 (84.0%) | +34.0 | (+10.1, +59.1) | **0.006\*** |
| Day 2 (Baseline: n=26) (Endline: n=24) |  |  |  |  |  |
| Dose | 13 (50.0%) | 23 (95.8%) | +45.8 | (+17.8, +66.8) | **0.001\*** |
| Frequency | 11 (42.3%) | 16 (66.7%) | +24.4 | (-4.4, +50.6) | 0.100 |
| Duration | 14 (53.8%) | 20 (83.3%) | +29.5 | (+6.4, +55.2) | **0.013\*** |
| **Paracetamol** |  |  |  |  |  |
| Day 0 (Baseline: n=78), (Endline: n=72) |  |  |  |  |  |
| Dose | 38 (48.7%) | 63 (87.5%) | +38.8 | (+24.4, +52.6) | **<0.001\*** |
| Frequency | 11 (14.1%) | 14 (19.4%) | +5.3 | (-8.1, +18.3) | 0.447 |
| Duration | 39 (50.0%) | 11 (15.3%) | -34.5 | (-49.4, -19.8) | **<0.001\*** |
| Day 1 (Baseline: n=84), (Endline: n=80) |  |  |  |  |  |
| Dose | 56 (66.7%) | 78 (97.5%) | +30.8 | (+19.1, +42.7) | **<0.001\*** |
| Frequency | 15 (17.9%) | 69 (86.3%) | +68.4 | (+56.9, +81.1) | **<0.001\*** |
| Duration | 54 (64.3%) | 73 (91.3%) | +27.1 | (+15.4, +39.4) | **<0.001\*** |
| Day 2 (Baseline: n=76), (Endline: n=76) |  |  |  |  |  |
| Dose | 44 (57.9%) | 75 (98.7%) | +40.8 | (+28.1, +53.5) | **<0.001\*** |
| Frequency | 12 (15.8%) | 20 (26.3%) | +10.5 | (-4.2, +25.2) | 0.162 |
| Duration | 28 (36.8%) | 31 (40.8%) | +4.0 | (-10.9, +18.9) | 0.600 |

RD, risk difference (endline proportion minus baseline proportion), expressed in percentage points; 95%
CI, 95% confidence interval; \* Significant at p-value < 0.05.

#### Baseline overall mean care score

At baseline, the summed percentage compliance across all 65 components was 3,036.0%. Divided this by the total number of components.

Baseline Mean Care Score = 3,036.0%/ 65 = 46.7%.

#### Endline overall mean care score

At endline, the summed percentage compliance across all standards was 3,970.6%, resulting in: Endline Mean Care Score = 3,970.6% / 65 = 61.1%.

The overall mean care score improved from 46.7% at baseline to 61.1% at endline, reflecting an absolute increase of +14.4% points in compliance with recommended postoperative care standards. Despite this improvement, the overall level of adherence remained low, indicating persistent gaps in the quality of postoperative monitoring and management.

## Discussion

This clinical audit demonstrated that targeted quality improvement interventions can improve selected aspects of postoperative care among mothers undergoing emergency caesarean section due to obstructed labour. Significant improvements were observed in prophylactic antibiotic administration, empiric antibiotic administration, analgesia administration, and selected aspects of bladder care following implementation of the audit recommendations. However, no significant improvements were observed in several critical standards, including vital sign monitoring, uterine tone and vaginal bleeding monitoring, and surgical site care. Although the overall mean care score increased from 46.7% at baseline to 61.1% at endline, adherence to recommended postoperative care standards remained suboptimal. These findings are consistent with previous obstetric audit studies that demonstrated the potential of clinical audits to improve quality of care while highlighting the influence of health system constraints on audit outcomes [9], [13].

The observed improvement in prophylactic antibiotic administration suggests that relatively simple system-level interventions can positively influence clinical practice. Similar findings were reported by Kayiga et al. who documented an increase in intravenous antibiotic administration following implementation of audit recommendations in the management of obstructed labour at Mulago National Referral Hospital [9]. Likewise, Akello et al. (2023) reported improved adherence to antibiotic administration protocols following a clinical audit on the management of preterm birth [8]. The improvement observed in the present study may be attributed to integrating antibiotic administration into routine theatre workflows and strengthening documentation through the hospital electronic medical records system. Nevertheless, the persistence of incomplete compliance indicates that barriers to consistent implementation remain and that additional interventions may be required to achieve optimal adherence. This observation supports findings by Kayiga et al. who noted that although audits improved quality of care, achievement of recommended standards remained challenging [9].

Compliance with empiric antibiotic administration also improved substantially, particularly for metronidazole, where high levels of adherence were achieved across multiple postoperative time points. These findings suggest that interventions targeting drug availability, medication dispensing processes, and documentation practices can significantly improve treatment compliance. However, the failure to sustain improvements in ceftriaxone administration beyond the immediate postoperative period highlights the challenges associated with maintaining adherence over time. Similar observations were reported by Mgaya et al. in Tanzania, where some improvements following audit interventions were modest and difficult to sustain [13]. The authors concluded that persistent gaps in standard care indicate ongoing opportunities for quality improvement if healthcare providers maintain commitment to implementing recommended practices. Our findings similarly emphasize the need for continuous monitoring, supervision, and reinforcement of postoperative care protocols.

The significant improvement observed in administration of postoperative analgesia demonstrates the potential effectiveness of patient-centred approaches in improving pain management. Encouraging mothers and caregivers to actively participate in reminding healthcare providers about scheduled medications may have contributed to improved compliance with prescribed analgesics. However, the decline in tramadol administration following the intervention illustrates the possibility of unintended consequences during quality improvement initiatives. This finding suggests that some healthcare providers may have interpreted the intervention as favouring self-administered non-opioid analgesics at the expense of other prescribed medications. Similar challenges have been highlighted by Kayiga et al. who reported that some standards failed to improve despite implementation of protocols because of inadequate clarity regarding expected practices [9]. The findings underscore the importance of clear and unambiguous communication during implementation of quality improvement interventions.

Bladder care demonstrated mixed results. Although documentation of urine output improved significantly, compliance with prolonged catheter management remained poor. The improvement in documentation may reflect increased awareness among healthcare providers following audit feedback and continuing medical education sessions. However, adherence to standards relating to catheter duration and discharge with a urethral catheter remained low. These findings differ from those reported by Mgaya et al., who found consistently high compliance with bladder catheterization practices before and after their audit intervention [13]. The discrepancy may be explained by differences in the audit standards used. Whereas Mgaya et al. evaluated the presence of catheterization, the current audit assessed adherence to a more stringent standard requiring maintenance of a urethral catheter for a specified duration among mothers with obstructed labour. This distinction highlights the importance of considering contextual and clinical differences when comparing audit outcomes across settings.

Despite improvements in some standards, vital sign monitoring remained consistently poor throughout both audit cycles. Similar findings have been reported in previous audits conducted in Uganda. Kayiga et al. observed that monitoring of maternal blood pressure and pulse rate showed little improvement following implementation of audit recommendations [9]. Mugyenyi et al. also reported suboptimal monitoring of vital signs among obstetric patients and attributed this to high patient volumes and limited staffing [14]. Effective monitoring is a fundamental component of postoperative care because it facilitates early identification of complications such as haemorrhage, sepsis, and hemodynamic instability. In the present study, the lack of improvement may reflect persistent systemic challenges, including high patient loads, workforce shortages, inadequate supervision, and limited availability of functional monitoring equipment. These findings suggest that educational interventions alone may be insufficient to improve monitoring practices in resource-constrained settings and that broader health system strengthening efforts are required.

Although the overall quality of postoperative care improved following implementation of the clinical audit, the magnitude of improvement was modest. The overall mean care score remained below acceptable levels, indicating that a substantial proportion of mothers continued to receive care that did not fully meet established standards. This finding contrasts with other studies which reported a decline in overall quality scores during re-audit [9], but is consistent with conclusions from [13] that successful implementation of audit recommendations depends on the availability of functional healthcare systems, including adequate staffing, supplies, equipment, teamwork, and supportive supervision. The persistence of suboptimal care despite targeted interventions suggests that sustainable improvements in postoperative management require not only audit and feedback mechanisms but also investments in health system capacity.

### Strengths and limitations

A major strength of this study was the use of a criterion-based clinical audit approach, which allowed objective assessment of adherence to evidence-based postoperative care standards and evaluation of changes following implementation of quality improvement interventions. The study also addressed an important evidence gap regarding postoperative management of women undergoing emergency caesarean section due to obstructed labour in a high-volume referral hospital.

However, several limitations should be considered when interpreting the findings. First, the study was conducted in a single referral hospital, which may limit generalizability to other healthcare settings. Second, some outcomes relied on documentation in patient records, and undocumented care may have resulted in underestimation of actual clinical practice. Third, the relatively short interval between implementation of interventions and re-audit may have limited the ability to assess long-term sustainability of improvements. Finally, persistent resource constraints, including shortages of monitoring equipment and high patient workloads, may have influenced adherence to some standards independently of the audit intervention.

## Conclusion

This clinical audit demonstrated improvements in selected aspects of postoperative care following emergency caesarean section due to obstructed labour, after implementation of targeted quality improvement interventions. Significant improvements were observed in prophylactic and empiric antibiotic administration, analgesia administration, and selected components of bladder care. However, deficiencies remained in vital-sign monitoring and other key aspects of postoperative management, indicating that improvements were non-uniform across all audited standards. These findings highlight the importance of combining clinical audit and feedback with strengthening of health system resources, staffing, equipment availability, and supportive supervision. Future quality improvement initiatives should focus on addressing these systemic barriers to ensure consistent adherence to evidence-based postoperative care standards and improve maternal outcomes.

## Data Availability

The data underlying this study contain potentially identifying and sensitive patient information (individual clinical case records of women who underwent emergency caesarean section due to obstructed labour). Public sharing of the complete dataset is restricted by the School of Medicine Research Ethics Committee (SOMREC) at Makerere University to protect patient privacy and confidentiality. However, the minimal de-identified dataset necessary to replicate the findings is available upon reasonable request from the corresponding author (Mwanje Derrick) or the ethics committee (SOMREC-2025-599)

## Acknowledgments

We thank the administrations of Kawempe National Referral Hospital and Makerere University for their support towards the completion of this study.

## Author contributions

**Conceptualization**: Mwanje Derrick, Kagawa Nantamu Mike, Nanzira Racheal Samantha, Byaruhanga Kayogoza Emmanuel, Igirimbabazi Ssebuddo Pious, Ononge Sam

**Data curation**: Mwanje Derrick, Igirimbabazi Ssebuddo Pious, Kagawa Nantamu Mike, Ononge Sam, Nanzira Racheal Samantha

**Formal analysis**: Mwanje Derrick

**Methodology**: Mwanje Derrick, Kagawa Nantamu Mike, Byaruhanga Kayogoza Emmanuel, Ononge Sam, Nanzira Racheal Samantha

**Validation**: Kagawa Nantamu Mike, Ononge Sam, Nanzira Racheal Samantha

**Writing– review & editing**: Mwanje Derrick, Kagawa Nantamu Mike, Ononge Sam, Nanzira Racheal Samantha.

